# Intermittent Theta-burst Transcranial Temporal Interference Stimulation focusing on the Putamen improves Symptom Severity in Parkinson’s Disease – A randomized controlled Trial

**DOI:** 10.64898/2026.02.05.26345627

**Authors:** Johannes Stalter, Heiko Stecher, Linda Bergmann, Pedro Arizpe-Gómez, Andreas Hein, André Aleman, Christoph S. Herrmann, Karsten Witt

**Affiliations:** Department of Neurology, Carl von Ossietzky University Oldenburg, Heiligengeisthöfe 4, 26121 Oldenburg, Germany; University Hospital of Neurology, Evangelical Hospital Oldenburg, Steinweg 13 – 17, 26122 Oldenburg, Germany; University of Groningen, Cognitive Neuroscience Center, University Medical Center Groningen, Hanzeplein 1, 9713 GZ Groningen, The Netherlands; Experimental Psychology Lab, Carl von Ossietzky University Oldenburg, Ammerländer Heerstraße 114 – 119, 26129 Oldenburg, Germany; Assistance Systems and Medical Device Technology, Carl von Ossietzky University Oldenburg, Ammerländer Heerstraße 114 – 119, 26129 Oldenburg, Germany; Faculty of Psychology and Neuroscience, Maastricht University, Universiteitssingle 40, 6229 ER Maastricht, The Netherlands; Center of Neurosensory Sciences, Carl von Ossietzky University Oldenburg, Ammerländer Heerstraße 114 – 119, 26129 Oldenburg, Germany

**Author notes:** Correspondence to: Johannes Stalter, MD, Dept. Of Neurology, University Oldenburg, Heiligengeisthöfe 4, 26121 Oldenburg, Lower Saxony, Germany. This study is published as a preprint under https://doi.org/10.64898/2026.02.05.26345627 and has been presented at the International Brain Stimulation Conference 2025.

**Keywords:** Parkinson’s disease, Neuromodulation, Temporal Interference Stimulation, Motor Learning, Deep Brain Stimulation

## Abstract

Transcranial temporal interference stimulation (tTIS) is a non-invasive approach for targeting deep brain regions such as the basal ganglia. This randomized, double-blind, crossover study examined whether intermittent theta-burst tTIS (iTBS-tTIS) applied to the right putamen reduces symptom severity in Parkinson’s disease (PD) and enhances motor learning and performance. Nineteen PD patients and 19 matched healthy controls underwent structural MRI and individualized electric field simulations. Primary outcome was the motor score of the left-sided MDS-UPDRS III; secondary outcomes included motor performance (alternating tapping task) and motor learning (sequential finger tapping task).

iTBS-tTIS significantly reduced MDS-UPDRS scores in PD patients, and improvement correlated with simulated electric field strength in the right putamen. However, no significant effects were observed for motor performance or motor learning in either group.

These results suggest that iTBS-tTIS may offer a promising non-invasive strategy to modulate deep brain structures and alleviate motor symptoms in PD.

## 1 Introduction

Transcranial temporal interference stimulation (tTIS) is a newly developed electrical technique for noninvasively stimulating deep brain regions without affecting the overlying cortex.^1^ This method tackles the issues of the depth-focality tradeoff of “conventional” non-invasive brain stimulation techniques. tTIS is performed by applying two very high-frequency oscillations with slightly different frequencies (e.g., 2 and 2.1 kHz; carrier frequency) and an overlap in the region of interest (ROI), such as the basal ganglia (BG).^1^ In this ROI, the currents interfere, resulting in a carrier frequency that is amplitude-modulated (AM) at the difference frequency of both frequencies (envelope frequency). This AM frequency can then interfere with neuronal activity in the target region. This indicates that the biological effects are mainly restricted to this specified region if a certain threshold is reached (Figure 4A). However, it is noteworthy that the brain is generally exposed to high carrier frequencies. Finite element modeling (FEM) has demonstrated that tTIS results in intracranial e-fields strong enough to modulate neuronal activity.^2^ Intracranial recordings in epilepsy patients have found tTIS induced e-fields in the hippocampus exceeding the threshold for modulating neuronal activity in the target region.^3^ For an overview of this technique, see recent literature.^4–7^ By calculating the optimal electrode montage together with the applied current, the target region can be individually optimized and steered without physically moving the electrodes. tTIS has been proven safe and effective for stimulating deep brain regions in humans.^8–10^

Parkinson’s disease (PD) is characterized by the loss of dopamine neurons in the substantia nigra (SN). As a result of this loss, a dysfunction in the neural loops connecting the BG with cortical areas occurs.^11^ Motor dysfunction is expressed by an increase in ß-oscillations with an anti-kinetic effect and a decrease in L-oscillations that diminish their prokinetic effects.^12–14^ In PD, these disturbances in the motor network can be attributed to altered oscillations in anatomically segregated pathways that are targeted by electrical stimulation as a treatment option.^15^ The rationale for using electrical stimulation to treat PD can best be described by the use of deep brain stimulation (DBS) in PD, which decreases ß-power in the subthalamic nucleus (STN) in parallel with an improvement in motor symptoms. However, this method is limited by its invasiveness.^12,16–18^ Therefore the idea of non-invasive stimulation techniques as a treatment option emerged, but have not been sufficiently effective in PD, possibly due to their low focal intensity or limited stimulation depth.^19,20^ Previous non-invasive brain stimulation methods, such as transcranial direct current stimulation (tDCS) and (repetitive) transcranial magnetic stimulation ((r)TMS) and ultrasound-based methods have been used in Parkinson’s disease for motor as well as non-motor symptoms.^21,22^ Studies with direct (electro)-physiological readouts such as fMRI or EEG could show effects not only at the stimulation side but also on a network-level.^23^ However, owing to the large variety of parameters and techniques, only a few results were replicated. This variety in methods and parameters holds huge potential for individualized treatment options. For a more comprehensive review, please see Bange et al. 2025.^21^ tTIS may overcome these disadvantages and can be applied with different stimulation settings, such as excitatory low frequencies, inhibitory high frequencies, and an intermittent theta burst (iTBS) protocol, which has been shown to elicit long-lasting excitatory features (e.g., plasticity induction) using different stimulation modalities, such as transcranial magnetic stimulation, ultrasound stimulation, or electric field stimulation. iTBS with 5 Hz is derived from hippocampal slices of mice models and the naturally occurring 5 Hz rhythm measured there.^24–26^ This stimulation pattern has been proven to effectively elicit long-lasting effects on a network-level and on a clinical-level (motor function and quality of life) in PD when applied with transcranial magnetic stimulation.^27,28^ tTIS therefore offers the potential to combine the positive effects of a cost-effective non-invasive therapy option with very little side-effects. Recently, it has been highlighted that there is a need for individualized electrical field simulation of non-invasive electrical stimulation to account for interindividual differences in anatomical variability, even more in pathologically altered brains.^29–33^ To account for this, the electrical fields produced by tTIS can be simulated on a single subject level.^29^

In this study, we applied iTBS-tTIS in a randomized, double-blinded crossover study design in PD and healthy controls. Given its excitatory effect of iTBS on neuronal tissue, we hypothesized that iTBS-tTIS applied to the right putamen would enhance neural communication at the entry point of the basal ganglia-thalamo-cortical loop system and reduce the severity of motor symptoms in PD patients. In an exploratory approach the strength of the simulated e-field in the right putamen is correlated with the clinical effect of the stimulation. To investigate whether healthy brain networks react differently compared to pathologically altered brains, in an additional exploratory approach, a healthy control group was included. This study aimed to assess the efficiency and feasibility of using iTBS-tTIS to target the putamen in patients with PD.

## 2 Results

### 2.1 Demographic and neuropsychological characteristics

Nineteen PD patients and 19 healthy seniors participated and completed the study between October 2023 and June 2024. No significant differences were observed between groups in age or gender (see Table 1). Global cognitive competence as measured by the MoCA showed a significantly lower score in the PD group (22.9±62.65 vs. 27.2±1.05, p<0.001), as well as in the paper pencil version of Symbol Digit Modality Test (SDMT) (PD 36.4 ± 11.50 vs. HC 47.0 ± 8.44, p=0.002). No participants screened were demented according to a MoCA Score < 18 points. BDI II scores also did not significantly differ between PD patients and HC. No participant was classified as depressed according to a BDI II score > 21 points. None of the participants stated to be a smoker and all participants were right-handed according to the *Edinburgh Handedness Scale.* The mean time period between the two stimulation sessions was 10.31 days (HC mean 8.1 min/max 6/16, PD mean 10.58 min/max 6/15) across both groups.

**Table 1:** Baseline and clinical characteristics.

|  | PD group | HC group | p-value |
| --- | --- | --- | --- |
| Age (years, mean, SD) | 64.70 (9.27) | 68.60 (6.26) | 0.09 |
| Sex (m/f) | 13/6 | 10/9 | 0.37 |
| Handedness (r/l) | 19/0 | 19/0 |  |
| Most affected side (right/left) | 8/11 | - |  |
| MoCA (mean, SD) | 22.90 (2.65) | 27.2 (1.05) | $< 0.001$ |
| SMDT (mean, SD) | 36.4 (11.50) | 47.0 (8.44) | 0.001 |
| BDI (mean, SD) | 7.78 (6.15) | 5.11 (4.18) | 0.245 |

Clinical data of the PD group
|  | N | Minimum | Maximum | Mean | Std. Dev. |
| --- | --- | --- | --- | --- | --- |
| UPDRS_I (all items) | 19 | 0 | 18 | 6.94 | 5.14 |
| UPDRS_II (all items) | 19 | 0 | 16 | 8.36 | 4.51 |
| UPDRS_III (all items) | 19 | 7 | 41 | 19.0 | 9.26 |
| UPDRS_IV (all items) | 19 | 0 | 8 | 1.26 | 2.077 |

SD = Standard deviation; PD = Parkinson's disease; HC = Healthy controls; m= male; f = female; MoCA = Montreal Cognitive Assessment; SMDT= Symbol Digit Modality Test; BDI = Becks Depression Inventory; MDS-UPDRS = Movement Disorder Society – Unified Parkinson's Diseases Rating Scale

Further clinical data of the PD patient group are given in Table 1. The MDS-UPDRS part III in total revealed a mean of 19.26 (± 9.26) points. With means at the first session of 19.26 (± 8.89) and at the second session of 17.42 (± 8.84) (p = 0.21), there was no significant difference between both stimulation sessions. The average Hoehn & Yahr stage was 2.3 (± 0.4).

Eleven patients (57.89%) showed the left side to be the dominantly affected side, while eight patients (42.11%) were more impaired on the right side. The mean disease duration was 6.26 years (± 4.43). For the LEDD a mean of 494.93 mg (± 309.76) was calculated. All participants were included in the following analyses.

### 2.2 Clinical Outcomes

#### 2.2.1 MDS-UPDRS

The change of the sum score of the analyzed (items 3.3, 3.4, 3.5, 3.6, 3.15, 3.16, 3.17, 3.18) MDS-UPDRS left sided items of the extremities between pre to during real stimulation and the pre to during sham condition revealed a significant greater improvement of the real stimulation with a moderate to large effect size (W = 30.5, p = 0.015, r = 0.49, Mann-Whitney-U test). Table 2 shows the changes for the analyzed MDS-UPDRS part III and the different sub-scores for bradykinesia (3.4, 3.5, 3.6), tremor (3.15, 3.16, 3.17, 3.18) and rigidity (3.3) of the left extremities. An analysis of the subscores revealed a significant reduction of the rigidity symptoms while the other subscores remained non-significant.

**Table 2.**
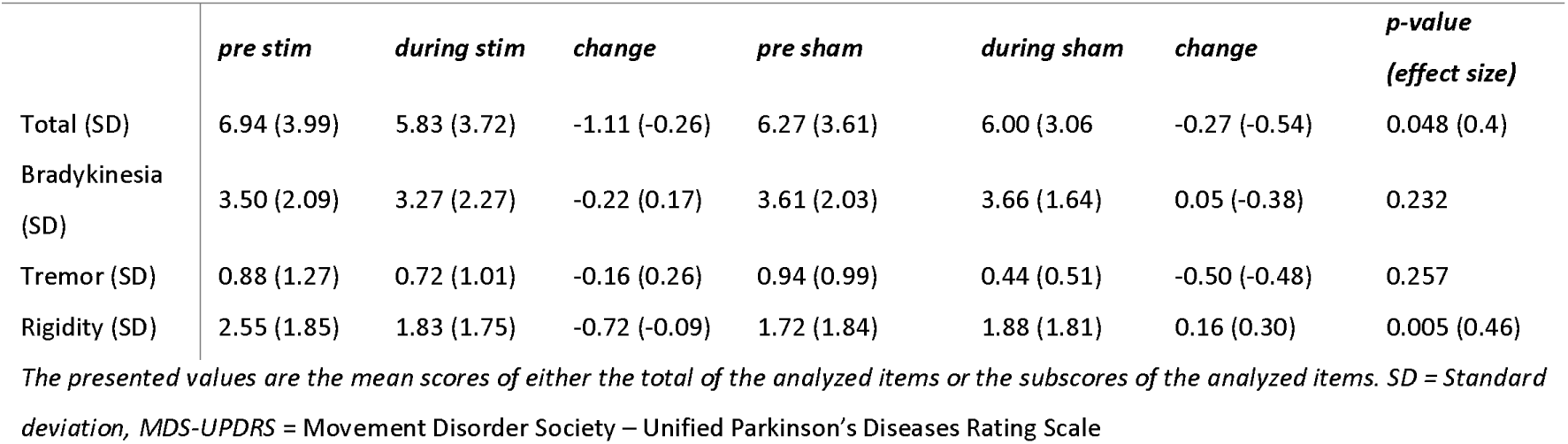
Changes of the left-sided MDS-UPDRS part III.

|  | *pre stim* | *during stim* | *change* | *pre sham* | *during sham* | *change* | *p-value*<br>*(effect size)* |
| --- | --- | --- | --- | --- | --- | --- | --- |
| Total (SD) | 6.94 (3.99) | 5.83 (3.72) | -1.11 (-0.26) | 6.27 (3.61) | 6.00 (3.06) | -0.27 (-0.54) | 0.048 (0.4) |
| Bradykinesia (SD) | 3.50 (2.09) | 3.27 (2.27) | -0.22 (0.17) | 3.61 (2.03) | 3.66 (1.64) | 0.05 (-0.38) | 0.232 |
| Tremor (SD) | 0.88 (1.27) | 0.72 (1.01) | -0.16 (0.26) | 0.94 (0.99) | 0.44 (0.51) | -0.50 (-0.48) | 0.257 |
| Rigidity (SD) | 2.55 (1.85) | 1.83 (1.75) | -0.72 (-0.09) | 1.72 (1.84) | 1.88 (1.81) | 0.16 (0.30) | 0.005 (0.46) |

*The presented values are the mean scores of either the total of the analyzed items or the subscores of the analyzed items. SD = Standard deviation, MDS-UPDRS = Movement Disorder Society – Unified Parkinson's Diseases Rating Scale*

#### 2.2.2 Simulation results

The calculation of the mean e-field strength in the right putamen showed a comparable e-field of both groups (PD: 0.226 V/m (±0.022) vs. HC: 0.242 V/m (± 0.031), p = 0.07, r = 0.43). One outlier (e-field = 0.36 V/m) was eliminated from the analysis. Pearson correlation analysis revealed a significant, moderate-to-strong negative correlation between the calculated e-field and improvement in motor function (p = 0.038, r = -0.49, Figure 1). We found no significant association between e-field intensity in the putamen and performance on the aTT and sFTT, nor did we find a significant association between e-field intensity in the STN and GPI and changes in the MDS-UPDRS-III, which suggests that these structures are unlikely to influence motor changes.

**Figure 1:**
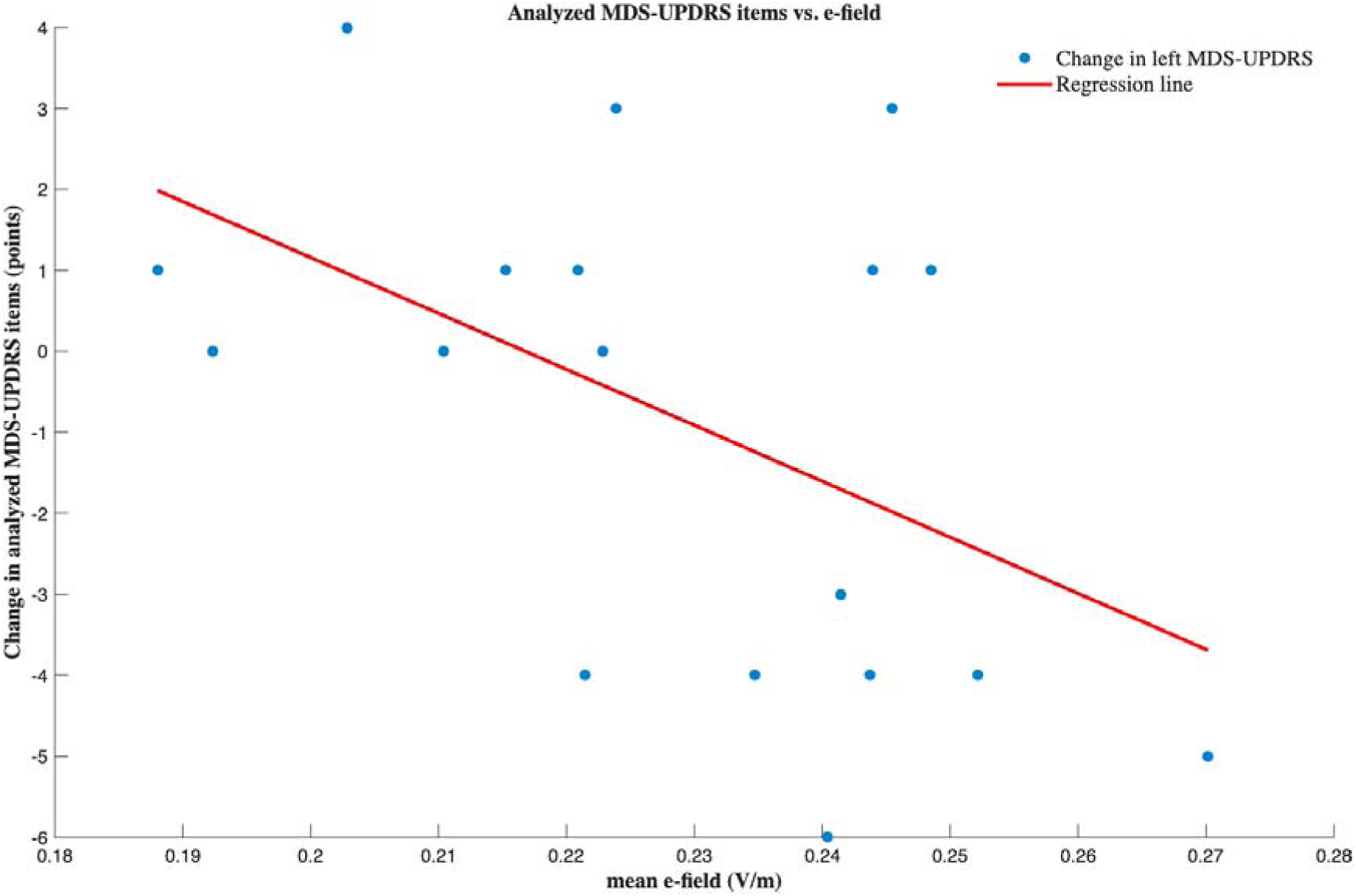
Correlation of the calculated change in the left-sided MDS-UPDRS and the simulated e-field of the right putamen. Blue points indicate individual patient data; the red line shows the regression line. There is a significant correlation between the mean e-field and the MDS-UPDRS. (MDS-UPDRS = Movement Disorder Society – Unified Parkinson’s Disease Rating Scale.)

### 2.3 Alternating Tapping Task

The linear mixed effects (LME) model applied to the aTT data showed significant effects for trial (chi^2^ = 13.987, p = 0.007), block (chi^2^ = 104.916, p < 0.001) and stim (chi^2^ = 64.322, p < 0.001) while group presented only a trend (chi^2^ = 3.087, p = 0.079). The interaction block x stim (chi^2^ = 50.825, p < 0.001), block x group (chi^2^ = 48.761, p < 0.001) and stim x group (chi^2^. = 51.066, p < 0.001) were significant. Bonferroni-adjusted post-hoc tests revealed no stable significant interaction for the tested variables (figure 2). Incorporating age as a covariate in the LME did not significantly affect the outcomes of the aTT.

**Figure 2:**
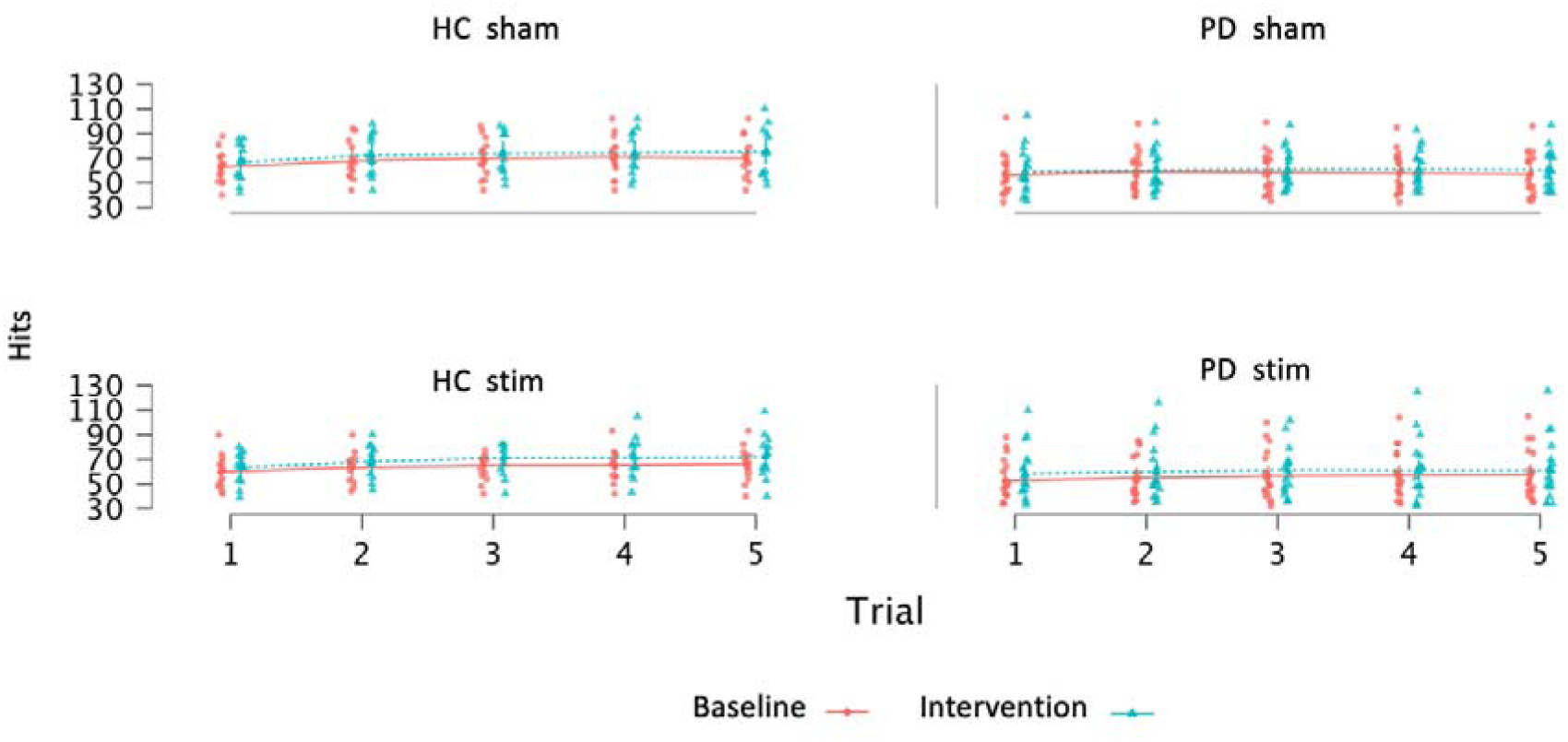
Hits are defined as key presses on two different buttons. There was a significant improvement between the baseline and tTIS in the control group. No significant change was observed in the Parkinson’s disease group. HC = Healthy controls; PD = Parkinson’s disease.

### 2.4 Sequential Finger Tapping Task

The linear mixed effect model (LME) showed significant effects of block (chi^2^ = 10.967, p < 0.001), stim (chi^2^ = 11.929, p < 0.001), Group (chi^2^ = 7.823, p = 0.005) and trial (chi^2^ = 31.304, p < 0.001). Across groups and stimulation conditions, accuracy improved from the first to the second block and over trials, indicating overall learning effects. Furthermore, healthy controls showed overall higher accuracy than patients with Parkinson’s disease.

Investigating the interactions, stim x group interaction was significant (chi^2^ = 10.342, p = 0.001), showing a greater negative influence of stimulation in the PD than in the HC group. Furthermore, the interaction stim x block x SDMT showed a significance (chi^2^ = 8.705, p = 0.013), indicating a negative association in SDMT and the performance in the task (sFTT). The Bonferroni adjusted post-hoc tests however did not reach a significant level for either group (figure 3). Incorporating age as a covariate in the LME did not significantly affect the outcomes of the sFTT.

**Figure 3:**
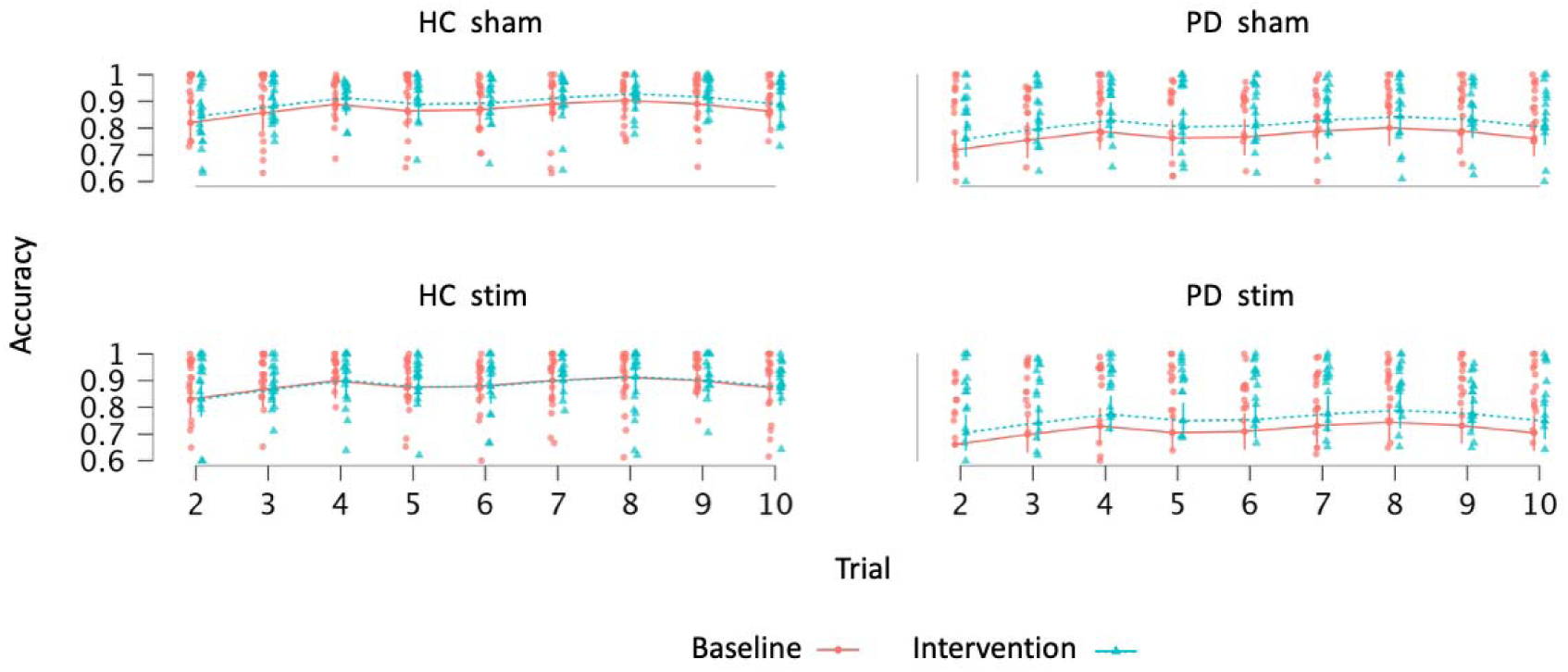
Results of the Sequential Finger Tapping Task. Upper panel: Sham sessions for both groups. Lower panel shows the data of the real stimulation tTIS sessions. Lines represent the marginal means while dots and triangles show individual data. Mean accuracy is calculated as the results of key presses within a correct sequence with all key presses of the trial. Again, No significant effects of real or sham stimulation were observed in either group. HC = Healthy controls; PD = Parkinson’s disease.

### 2.5 Side effects of non-invasive transcranial temporal interference stimulation

No severe side effects were reported in any group for any condition. Table 3 shows the reported side effects and how often they were reported. No individual side effect occurred significantly more frequently during stimulation than sham, as assessed by Fisher’s exact tests with FDR correction (p > 0.05). Consistent with this finding, the overall frequency of reported side effects did not differ between conditions (p>0.374).

**Table 3:** Reported side effects during and after the transcranial temporal interference stimulation.

| Symptom | Total_count | Ratio sham condition | Ratio stim condition | N_sham | N_stim | p_fisher |
| --- | --- | --- | --- | --- | --- | --- |
| Headache | 4 | 0.75 | 0.25 | 3 | 1 | 0.603 |
| Neck pain | 3 | 0.33 | 0.66 | 1 | 2 | 1 |
| Local pain | 2 | 0.5 | 0.5 | 1 | 1 | 1 |
| Dysesthesia | 5 | 0.2 | 0.8 | 1 | 4 | 0.339 |
| Burning sensation | 4 | 0.5 | 0.5 | 2 | 2 | 1 |
| Local warming | 4 | 0.25 | 0.75 | 1 | 3 | 0.603 |
| Redness | 13 | 0.46 | 0.53 | 6 | 7 | 1 |
| Fatigue | 19 | 0.47 | 0.52 | 9 | 10 | 1 |
| Concentration problems | 6 | 0.33 | 0.66 | 2 | 4 | 0.659 |
| Mood changes | 1 | 0 | 1 | 0 | 1 | 1 |

## 3 Discussion

In this study, iTBS-tTIS was combined with clinical examinations and motor tasks in a PD group and a control group. A short stimulation targeting the putamen improved the total left-sided MDS-UPDRS part III score assessed during stimulation as a clinical marker of symptom severity. Stimulation-induced changes in MDS-UPDRS part III correlated significantly with the simulated e-field in the right putamen. tTIS had no significant effect on motor-learning behavior or tapping behavior, neither in the HC nor in the PD group. No severe adverse side-effects were observed.

Baseline characteristics showed, in accordance with the disease, a well-matched selection of participants, with the PD data suggesting a clinically mildly affected group with moderate LEDD and moderate disease duration. The results of SDMT and MoCA showed an impairment of the processing speed and general cognitive function as expected in PD. Since cognitive processing speed can influence motor speed, especially in sequential finger-tapping tasks in Parkinson’s disease, these results are in line with the expected outcomes. Even though the tasks were explained in person, training trials were implemented for each task, specifically in the more cognitively demanding sequential finger tapping task, the cognitive component of the task was impaired in the patient group in the sFTT. Since patients with PD demonstrated relatively high accuracy scores, this impairment might have been partly counterbalanced by an accuracy-speed trade-off in this cohort. Nevertheless, these impairments in the PD group should be kept in mind when interpreting the results.

Mild adverse side effect that occurred were mostly of very short duration and lasted the longest until a few minutes after stimulation. The distribution between sham and stim condition was rather even, given the small number of reported cases.

We were able to enhance the performance of PD patients in MDS-UPDRS part III by using high gamma frequency with iTBS pattern over a short period of time. When correlating the improvement in total left-sided MDS-UPDRS part III with the simulated e-field in the ROI, a significant correlation could be shown, i.e. higher simulated e-fields were shown in participants with greater improvement under stimulation. This may provide a causal connection between the experimental stimulation and the clinical effects during the stimulation on patients and widens the knowledge and therapeutic potentials of non-invasive deep brain stimulation.^34^ Since a responder/non-responder split is visible in the data, with some patients even worsening under stimulation, this highlights the usefulness of the simulation and should be further explored in future studies. Another interpretation would be that low e-fields in the target region are an indirect indication of higher e-fields in adjacent region with different, e.g. negative impact on the motor behavior of participants. Furthermore, additional analysis indicates that subthreshold stimulation of the STN and GPi cannot account for the observed improvement in motor performance (supp. Table 1). Additionally, no correlation was found between electric field strength and its effect on sFTT and aTT.

One of the hallmarks of PD is the impaired movement, especially a slowing down, a reduction of the movement and irregularities in timing and a sequence effect (progressive reduction in velocity/amplitude over time) as well as a muscle stiffness (rigidity) as measured in the MDS-UPDRS part III.^35,36,37^ Typically, these parameters are decreasing more over the examination time. Dopaminergic dysfunction in PD is particularly pronounced in the SN, pars compacta where neurons are projecting into the putamen.^38^ ^39^ In the pathophysiological model of PD, due to the degeneration of dopaminergic neurons in the SN the inhibiting effect of the putamen to the GPi is decreased. In parallel, the inhibition of the STN decreases. These two mechanisms lead to an extensive excitation of the GPi, which in turn functions as the “exit nucleus” of the BG loop.^40,41^ Conventional DBS, which targets the STN or GPi, uses stimulation signals that disturb pathologically altered neuronal activity.^16^ It has been assumed that DBS of the STN or GPi has multiple effects, including a decrease in activity, reduction of acinetic beta-synchronization, a hallmark of PD, and desynchronization of network communication in the cortico-basal ganglia-thalamic-cortical loop system.^12,16,42^ This, in turn, leads to a normalization of the communication between the GPi and the ventral-lateral thalamic nucleus with its impact on the motor cortex, that is, changing the output of the BG loop. By choosing the “entrance” of the BG loop, that is, the putamen as the target region, our hypothesis points to the neuroplastic effects of an enhanced “entrance” signal from the cortex to the BG. By facilitating the effects of the putamen, the direct pathway, which promotes movement, is shaped, whereas the indirect pathway, which inhibits movement, may be further suppressed. ^43^ Therefore, in theory, using the putamen as a stimulation region not only targets the pathologically altered neuronal activity, like conventional DBS, but also may enable the BG system to diminish the pathologically disbalance between inhibitory and excitatory pathways.

Huang et al. showed that the effect of iTBS on the central nervous system emerges right after starting the stimulation.^44^ In addition, it is thought that not only pathological frequencies but also pathological neuronal firing patterns are phenomena closely connected with PD.^45^ Therefore it can be assumed that changing the entrance of the BG loop into a more physiological way, the effects on the following structures are not only changing the firing rate but also modulate the firing pattern. This is of special interest when considering the other loops of the BG, namely the limbic and the cognitive loop.

By improving symptoms in a certain patient cohort in a moderate disease stage with tTIS, it opens this research line as first steps towards a therapy approach for patients in earlier stages of the disease even before motor complications occur. However, the here presented data hold a certain inhomogeneity in its interpretation und therefore needs further validation before being translated into clinical practice. The improvement of the MDS-UPDRS part III during stimulation in Parkinson’s disease using tTIS in the region of the putamen with a plasticity modulating stimulation paradigm can be therefore interpreted as a first proof of feasibility. Yet, this study infers the effects to be plasticity induced from the chosen stimulation pattern. While the reported data prove an effect, the explanation that this is caused by plasticity changes remains hypothetical.

Only few studies investigated the effects of tTIS in Parkinson’s disease.^46–51^ Those other studies chose a stimulation paradigm closely related to the paradigm of conventional DBS, using tTIS to disrupt neuronal firing and therefore improve motor symptoms in Parkinson’s disease. By using an intermitted theta burst approach, we used neuronal excitatory and plasticity effects to modulate PD symptoms. Using an individualized approach to simulate the electrical fields for each individual, not only were individual differences taken into account, but also differences related to the pathological changes, such as atrophy with a consecutive larger cerebrospinal fluid volume with higher shunting effects, during the course of the neurodegenerative disease. Interestingly, there are no significant differences between the mean values when both groups are compared. Since a threshold of 0.2 V/m is thought to be necessary to sufficiently stimulate neurons on a subthreshold level, the individual e-field data for both groups show the potential of tTIS to stimulate deep brain regions.^52,53^ The comparability of both groups regarding the individualized e-fields might be explained by the short disease duration of the PD group. To our knowledge, no study investigated this so far and this finding adds valuable insights into the question of how individualized tTIS should be performed in a clinical setting. On the other hand, since a deterioration in the MDS-UPDRS part III to some degree took place during the sham stimulation, this may partly explain the effects measured in the clinical outcome. Possible causes are motor fluctuations - which are well known in PD - fatigue during the study sessions, medication or order effects as well as regression towards the mean. By randomizing the study design while the order within the sessions remained the same, we tried to counter-balance the effects of fatigue and order effects. The time of the study sessions were randomly placed during the day to avoid medication or circadian effects distorting the results of the measurements. Nevertheless, an influence of these factors cannot be completely excluded.

In this study, no enhancement in motor learning behavior was observed in either the control group or patients with Parkinson’s disease (PD). The absence of improvement in the PD group may be attributed to a theory proposed by Seidler et al., which suggests that learned motor behavior must be translated into movement patterns and execution.^54^ This process is decelerated in PD, potentially leading to a discrepancy between motor learning and motor performance. Consequently, if motor execution is compromised, motor learning may occur but remain unexecuted, a phenomenon known as learning-performance dissociation. Given that motor execution is impaired in Parkinson’s disease, this may partially explain the lack of improvement. Additionally, a strong placebo effect, common in PD, may reduce the potential effects of stimulation. Beyond these disease-specific factors, other potential explanations should be considered for both groups. For tTIS, no established dose-dependency exists, which may relate to the duration of stimulation. Moreover, a ceiling effect may have limited the magnitude of stimulation changes, preventing significant differences between groups or conditions from being detected.

Like the motor learning behavior, the motor performance showed no significant effect in any of the groups after post-hoc analysis. Considering the similar pattern across all conditions, it seems likely that there was a strong ceiling effect in the behavioral data. Given the relatively mild disease stage of the PD cohort, proximal movements may have been insufficiently sensitive to subtle changes in bradykinesia. This can also partly explain the difference between the clinical and the behavioral results. In addition, the limitations for the results of the sFTT apply to this task as well. Previous studies have consistently demonstrated the effects of tTIS when applied during a learning task. This is based on the assumption that iTBS enhances neuroplasticity during skill acquisition, leading to improved performance.^8,55^ Furthermore it is thought that tTIS does not depolarize neurons, thus producing facilitation-like oscillatory network effects. Improvements in the UPDRS-III, a more static measure in which characteristics such as rigidity and tremor were examined without motor conditioning, suggest that the previously performed motor tests (SFTT and aFTT) may have facilitated this conditioning. However, because the UPDRS-III was always assessed after the motor tests to maximize stimulation time, a suitable way to test this hypothesis would be to compare the UPDRS-III score taken before the motor tests under stimulation.

There was a certain heterogeneity in the PD group, mostly regarding the MDS-UPDRS part III and the affected side, which might have limited the effects in the present study, since the non-dominant side was stimulated. We assessed the left hand because the serial-sequence motor task was performed with the non-dominant hand. Previous studies have shown that learning in a motor learning task can be better assessed in the non-dominant hand because skill acquisition reaches a plateau more quickly in the dominant hand.^56^ In addition, stimulation took place in medication-ON, which might have masked stimulation effects, but in general, it indicates that a stimulation effect is strong enough to be demonstrated in an ON-medication condition. The randomization of sham and real conditions was chosen to control for slight fluctuations in medication levels that may occur throughout the day. A relatively short stimulation duration as well as low amplitude may be considered as stimulation-related limitations. We were able to prove the feasibility and short-term clinical effects of iTBS-tTIS leading to its investigation as a potential treatment option. Therefore, this cost-effective and safe method opens a new field of neuro-enhancement and techniques to understand cognitive functioning in pathologically altered states in deeper brain regions. Together with the clinically measured improvement in MDS-UPDRS part III, it can be interpreted as a proof for its feasibility and its potential as a new therapeutical approach. Since no severe adverse side-effects were reported in our study, which is in line with other experiments, one could think of higher stimulation currents to further improve the effects of non-invasive deep brain stimulation.^9^ Another limitation of the present study is the absence of physiological measures. To further enhance the understanding of the underlying mechanisms of iTBS-tTIS, future studies should include direct physiological measures, such as fMRI, EEG, or intracranial lead recordings. Since no severe adverse side-effects were reported in our study, which is in line with other experiments, one could think of higher stimulation currents to further improve the effects of non-invasive deep brain stimulation.^9^ Another limitation of the present study is the absence of physiological measures. To further enhance the understanding of the underlying mechanisms of iTBS-tTIS, future studies should include direct physiological measures, such as fMRI, EEG, or intracranial lead recordings.

In future studies, larger groups of more severely affected patients should be investigated to also allow for subgroup analyses, e.g. cognitive or motivational factors, effects of predominant clinically affected side and symptom severity that drive results. Additionally, investigations regarding the dose-dependency, potential after-effects and negative effects on clinical parameters should take place, not only in a PD study sample but also in healthy participants.

## Materials and methods

### 3.1 Participants

Patients were recruited from the outpatient clinic at the University Hospital Oldenburg, Germany. Inclusion criteria were a clinical diagnosis of PD according to MDS-criteria.^57^ Healthy participants were recruited via newspapers and matched by age and sex. All participants were right-handed (according to *Edinburgh Handedness Scale*), without a history of neurological or psychiatric diseases (except for PD in the PD group). Contraindications for MRI or electrical neurostimulation had to be absent. Further exclusion criteria were centrally acting medication (except for PD specific medication in the PD group) and severe cognitive deficits, defined as a Montreal Cognitive Assessment (MoCA) sum score of < 18 points in the PD and in the HC group.^58^ The range of the MoCA to 18 points addresses the fact that mild cognitive changes are frequently accompanying PD.^59^ The test material was presented, including simple task instructions, pre-test training, and the use of tests that minimized working memory load. All participants gave written informed consent before inclusion in the study. The study was conducted according to the Declaration of Helsinki, was approved by the local ethics committee and preregistered in the German Registry of Clinical Trials (DRKS00030841, date of registration 1. April 2023). The study was preregistered additionally on *Open Science Framework* (OSF, https://osf.io/u7sw5).

### 3.2 Study design

The study was conducted in a randomized, double-blind cross-over design. PD patients were examined in medication-ON. Randomization was done using a custom-written Matlab script, interventions were automatically assigned according to the randomization script. Neither the examiner nor the participant was able to unblind the intervention. The clinical assessment of the MDS-UPDRS part III was performed by a clinical neurologist trained in movement disorders from the outpatient clinic of the university hospital Oldenburg. The assessing neurologist entered the examination room after the stimulation setting was finished and the stimulation was running. The stimulation setup for sham and the actual stimulation was identical, and no conversation was allowed between the examiner and the patient (besides test instructions). The questionnaire about potential side-effects were filled in by the participants without the examiner and the patients were not known to the neurologist. In the first session, a detailed clinical examination took place, and MRI data were obtained for all participants. The second session contained a baseline measurement of all experimental motor tasks described below, followed by the same order of tasks during the intervention. After stimulation a questionnaire about the adverse effects was given to participants in each session. The third session was equivalent to the second session, with the corresponding condition (Figure 4C). Between sessions, a wash-out-phase of approximately ten days was established.

**Figure 4:**
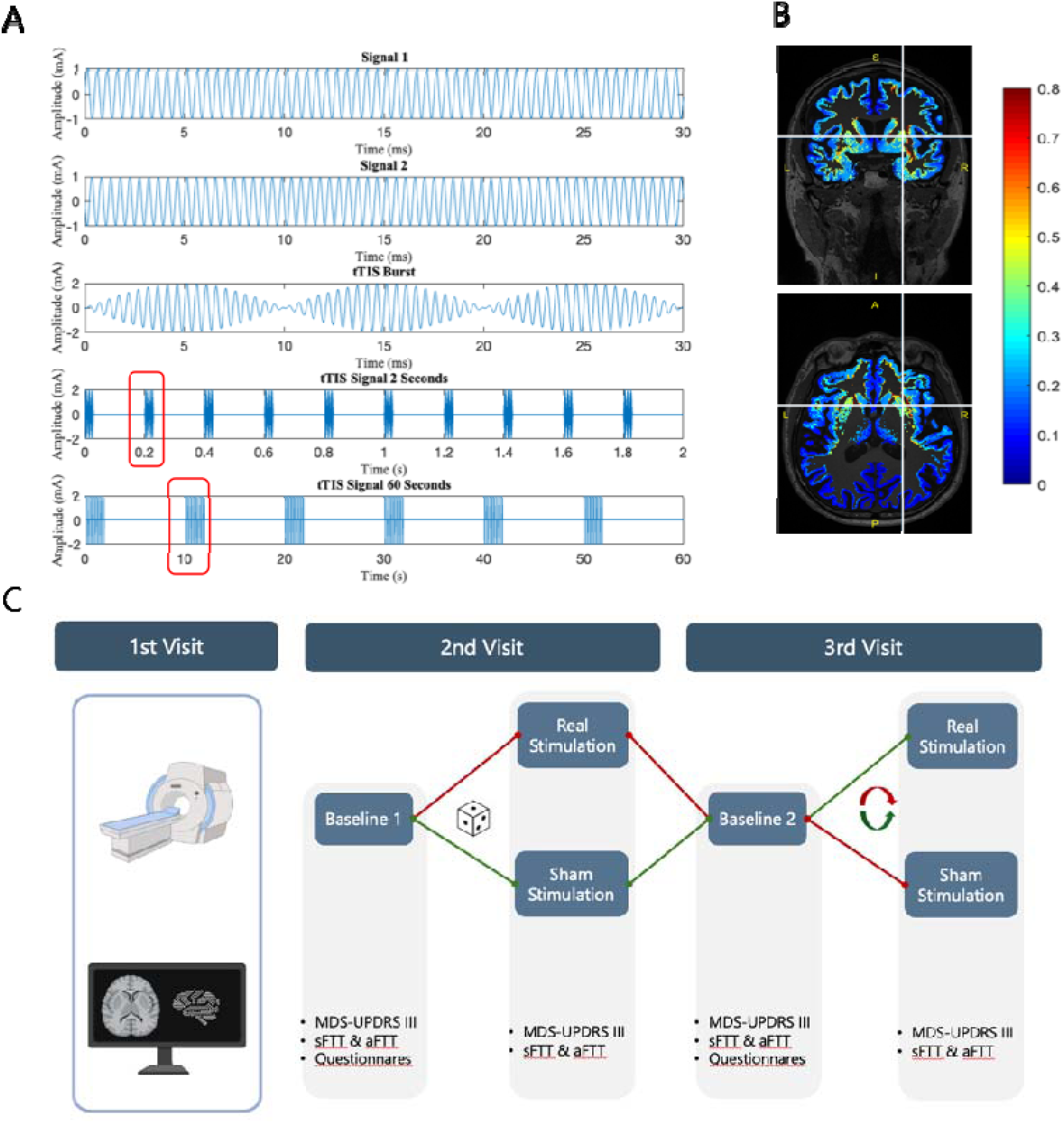
A) The first two rows illustrate two high-frequency signals (2 kHz and 2.1 kHz). The third row presents their combination, resulting in an amplitude-modulated tTIS signal characterized by three distinct peaks. These peaks are highlighted in red in the fourth row, which depicts ten trains of three peaks each over a two-second period. The fifth row illustrates the intermittent theta-burst pattern, where each two-second train is followed by an eight-second pause. This pattern repeats throughout the stimulation, with a 30-second ramp-up and ramp-down period at the beginning and end of the stimulation, respectively. For illustration purposes, the signal is shown as zero whenever no amplitude modulation occurs. During these times, both signals were oscillating at 2 kHz. (B) Simulation of transcranial temporal interference stimulation using electrode placements at F5/C5 and F6/C6, with a peak-to-peak current of 2 mA for each electrode pair. The color bar indicates the voltage gradient (V/m), representing the current reaching deep brain regions. The crosshairs mark the putamen. This simulation was generated using the MATLAB © toolbox SimNIBS 4.0 (Saturnino et al., 2019). A transversal section (upper figure) and a sagittal section (lower figure) are shown. (C) Study protocol: All participants were randomized (dice symbol) and double-blinded for assignment to the two study arms in a cross-over study design. (MRI = Magnetic Resonance Imaging; MDS-UPDRS III = Movement Disorder Society – Unified Parkinson’s Disease Rating Scale motor part; sFTT = Sequential Finger Tapping Task, aTT = Alternating Tapping Task).

### 3.3 MRI data

Structural imaging was acquired using a 3.0 Tesla MRI system (Siemens Magnetom Prisma, Syngo MR E11, Siemens Medical Solutions, Erlangen, Germany) using a 64-channel head–neck-coil. A T1-weighted Magnetization Prepared Rapid Gradient Echo Sequence (MPRAGE) was obtained together with a T2-weighted scan and a fluid-attenuated inversion recovery (FLAIR) sequence.

### 3.4 Simulation

To simulate stimulation-effects, SimNIBS 4.1 was used.^1,60^ Individual scans (T1 and T2) were segmented, and conductivity of tissue type was assigned. Using these meshes, simulation on a single-subject level was computed. Simulations were run with an injected current of 2 mA peak-to-peak for electrode pairs positioned at F5-F6/CP5-CP6. Using the melb-atlas^61^, we simulated the electrical field in the right putamen adapted to the individual neuroanatomy in the subject space of each participant (Figure 4B). Simulation was performed post-hoc only and therefore had no influence on individual stimulation parameters.

### 3.5 Stimulation

The stimulation was applied by two pairs of electrodes placed at F5-F6/CP5-CP6 (according to the international 10-10 system) attached to the scalp using ten20-paste (Weaver & Co, Aurora, CO, USA) with impedance below 10 kΩ. The right putamen was chosen since all participants self-reported being right-handed, e.g. the non-dominant side was assessed. We chose the intermittent theta-burst pattern as described by Wessel et al.^8^ This pattern has already been proven to modulate behavioral outcomes when applied to the BG. For stimulation, 5×5cm rubber electrodes were used with a current-intensity of 2 mA (peak-to-peak) since this is below the recommended maximum current density in skin of 0.1 mA/cm^2^ (= 2.5 mA zero-to-peak for 25cm^2^ electrodes).^10^

Stimulations started and ended with a ramp-up and a ramp-down of 30 seconds. The stimulation signal consisted of two sinusoidal currents. One was always oscillating at 2 kHz, the other was switched between 2 and 2.1 kHz. If the two frequencies were different, this resulted in an amplitude-modulated signal with a carrier frequency of 2.05 kHz (mean of the two frequencies) and a modulation frequency of 100 Hz (difference of the two frequencies). The switching of the frequency of the second current was chosen to result in periods of 30 ms – each containing three cycles of the 100 Hz amplitude-modulated oscillations (Figure 4A) targeting the putamen, followed by 170 ms with both signals oscillating at 2 kHz. This pattern is repeated over two seconds followed by a period with both signals oscillating at 2 kHz for eight seconds. This way, we mimicked a so-called Theta-Burst stimulation (5 Hz).^8^ The sham stimulation consisted of two signals both with a frequency of 2 kHz without frequency changes. The duration of stimulation was 45 minutes in total per session, separated in three blocks of 10, 20 and 15 minutes. The stimulation signals were created with a Matlab (V*ersion R2022b*. Natick, Massachusetts: The MathWorks Inc.) script and were converted to voltage signals via a digital-to-analog converter (Ni-USB 6251, National Instruments, Austin, TX, USA). The voltage signal was delivered by two battery-driven, galvanically isolated constant current stimulators (Advanced DC Stimulator Plus, Neuroconn, Ilmenau, Germany).

### 3.6 Clinical and Motor Assessments and Outcomes

The primary objective was to investigate the effects of iTBI-tTIS on motor target variables. We considered the change in MDS-UPDRS Part III on the left-sided extremities to be the primary endpoint. Secondary endpoints include alternating tapping behavior evaluating proximal arm movements and finger sequence learning using the left hand.^8^

#### 4.6.1 Clinical Assessment

For a clinical characterization, the MDS-UPDRS was used that included cognitive and affective domains (part I), management of activities of daily living (part II) and a detailed motor examination (part III) and motor complication such as dyskinesias and motor fluctuations (part IV).^57^ The total score can reach a maximum of 132 points with higher scores indicating higher PD symptom severity. For each patient, the levodopa equivalent daily dosage (LEDD) was calculated according to the formulars of Joost et al. 2023.^62^ The Montral Cognitive Assessment (MoCA) is used to assess the overall cognitive abilities over several domains.^63, 64^ In addition we measured cognitive processing speed using the Symbol Digit Modalities Test (SDMT).^65^ To screen for clinical relevant depression, we used the Beck Depression Inventory II, where a cut-off of 21 points indicates a clinical manifest depression.^66^ ^A^ll participants filled in the Edinburgh Handedness.^67^ The side-effects of the stimulation were recorded based on structured self-reports of the participants. The severity of the experienced sensations were asked on a scale from 0 to 4. After each session, the questionnaire had to be filled in.

#### 4.6.2 Motor Assessments and Outcome Parameters

The primary outcome of this study was the change in parkinsonian symptoms of the left extremities, defined as change in the left sided items of the MDS-UPDRS part III against the sham effect in PD. The left side was chosen, as the right putamen was targeted with the stimulation montage.

The secondary outcome parameters focused on the amplitude and speed of movements of proximal sections of the left extremity, as well as motor learning. Performance of these motor tasks was measured at baseline and under real or sham stimulation condition. The alternating tapping task (aTT) measures the motor performance by counting the number of key presses that can be performed in five trials of 30 seconds intersected by four 30 seconds of rest periods. The keys for this task were 30 centimeters apart. The outcome of the aTT is the total number of key taps in 30 seconds across five alternative tapping runs, analyzed using a repeated measurement design. The sequential finger tapping task (sFTT) was used to assesses motor skill learning.^56^ Participants were required to press four numeric keys on a computer keyboard with the fingers two to five of their left hand repeating a given five-element sequence (e.g. “4-2-3-1-2”, corresponding to the order “little finger– middle finger – ring finger – index finger – middle finger”) “as quickly and as accurately as possible” for a period of 30 seconds. Four different sequences were used in a counterbalanced order across the experimental runs. The numeric sequence is displayed on the screen in front of the participants at all times to minimize the working memory component of the task. Each block consisted of ten 30-second trails of sequential finger tapping followed a 30-second rest period. We selected an accuracy measure based on the quotient of correct finger taps in the given sequence divided by the total number of keystrokes achieved in 30 seconds. These quotients were then used for further statistical analysis.

### 3.7 Statistics

The data was analyzed and visualized using JASP Team (2025) (JASP [Version 0.95.4] and Matlab (V*ersion R2022b*. Natick, Massachusetts: The MathWorks Inc.). There are no previous studies that have estimated the effect size of iTB-tTIS stimulation in the putamen on motor outcomes (UPDRS-III). As an approximation, we based our calculations on the effect sizes for motor learning in healthy older adults (effect size = 1.19) and selected a power of 0.9, which resulted in a sample size of 18 study participants per group.^8^ To analyze demographic data, a Wilcoxon rank test was used for non-normally distributed values. Motor performance and motor learning was assessed with a linear mixed effects (LME) model, the variables group, condition, block and trial were used as fixed effects, the subject factor was inserted into the model as a random intercept. The left-sided items of the MDS-UPDRS part III were analyzed by calculating the difference between baseline and sham or baseline and stim condition. Wilcoxon rank test was used to compare those values.

## 4 Data availability

The data that support the findings of this study and the used codes are available from the corresponding author, upon reasonable request.

## 6 Author contributions

Conceptualization: JS, AA, CSH, KW; Data curation: JS; Formal analysis: JS, HS, LB, PAG, AH, CSH, KW; Funding acquisition: JS, AA, CSH, KW; Methodology: JS; HS, LB, PAG, AH, AA, CSH, KW; Project administration: JS; Resources: JS; AA, CSH, KW; Supervision: AA, Ah, CSH, KW; Writing: JS, CSH, KW; Writing editing: HS, LB, PAG, AA, CSH, KW

## Data Availability

All data produced in the present study are available upon reasonable request to the authors

## 9 Supplements

### 9.1 Calculated e-fields in the target region and adjacent regions

**Supp. table 1:**
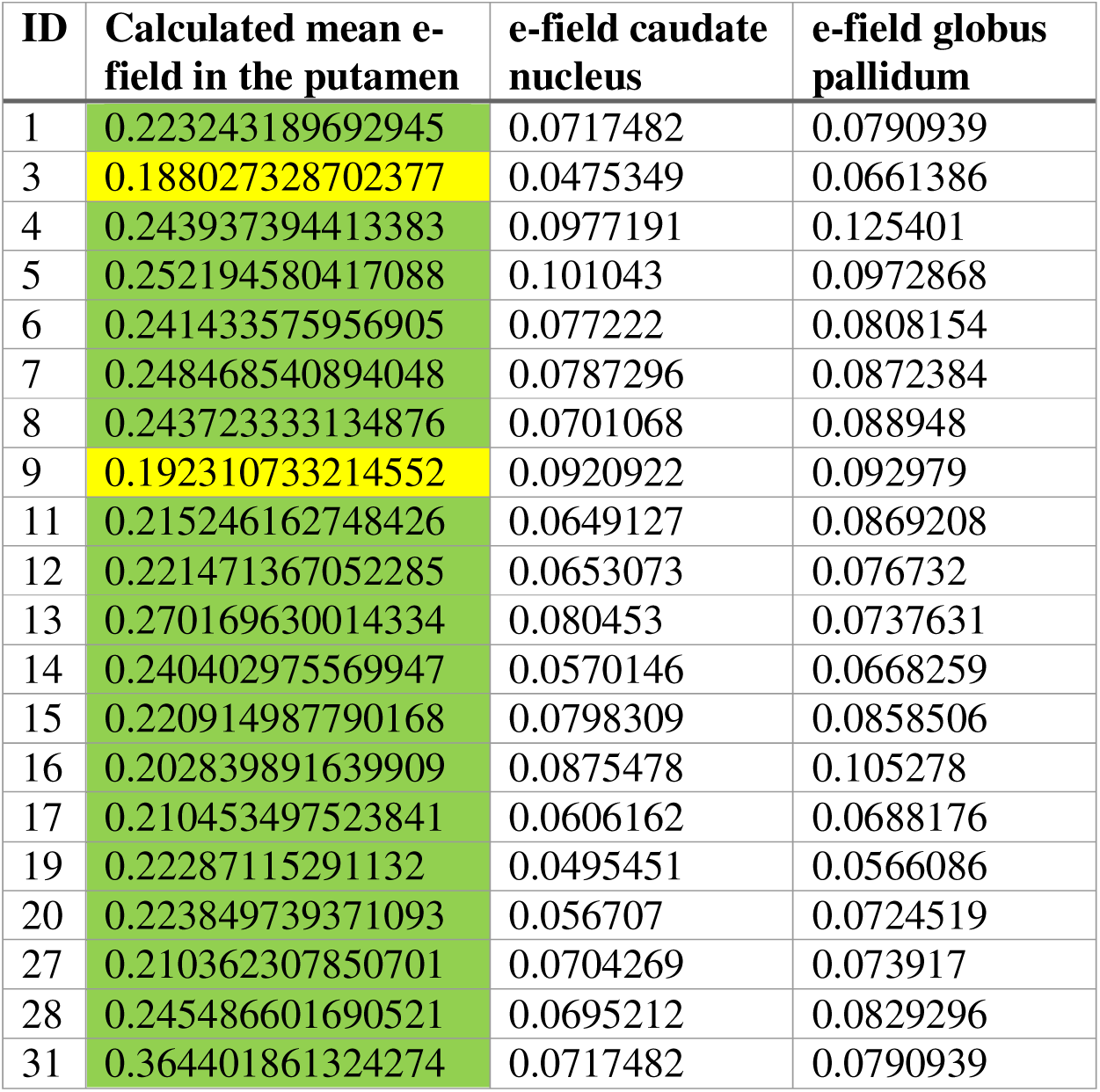
Detailed mean e-field summary of the PD group and indication if the threshold of > .2 mV/m was reached (green).

